# Evaluating the impact of a community-engagement intervention on the uptake of childhood vaccines in England: A synthetic control analysis

**DOI:** 10.64898/2026.05.01.26352232

**Authors:** Mohammed Sherif Amin, Xingna Zhang, Mark A. Green, Dawn Holford, Charlotte Hemingway, Amina Ismail, Nour Essale, Vicki Doyle, Miriam Taegtmeyer, Daniel Hungerford

**Affiliations:** Department of Clinical Infection Microbiology and Immunology, University of Liverpool, Liverpool, UK; Public Health, Policy and Systems (PHPS), University of Liverpool, Liverpool, UK; Civic Health Innovation Labs (CHIL), University of Liverpool, Liverpool, UK; Department of Geography and Planning, University of Liverpool, Liverpool, UK; School of Psychological Science, University of Bristol, Bristol, UK; Department of International Public Health, Liverpool School of Tropical Medicine, Liverpool, UK; Capacity Development International, Liverpool, UK; Department of Clinical Sciences, Liverpool School of Tropical Medicine, Liverpool, UK; NIHR Health Protection Research Unit in Emerging and Zoonotic Infections, University of Liverpool, Liverpool, UK

**Keywords:** Vaccination, Paediatrics, Epidemiology, Public health, Primary Prevention, inequalities, quasi-experimental

## Abstract

**Objective:** To evaluate the impact of equity-focused community-engagement initiatives on the uptake of five routine childhood vaccinations.

**Design:** Quasi-experimental study within a synthetic control analysis framework.

**Setting:** Primary care in England between April 2019 and March 2025. Childhood vaccination data were obtained from the Cover of Vaccination Evaluated Rapidly (COVER) programme.

**Intervention:** The Health Equity Liverpool Project (HELP) is a community-engagement vaccination initiative implemented between October 2023 and June 2024 across nine sites in central and north Liverpool. Activities were co-developed with local partners and delivered in neighbourhoods with persistently low childhood vaccine coverage. Intervention practices were defined as those located within 1 km of HELP delivery sites (n=19). A weighted combination of non-intervention practices across England (n=5826) was used to construct a synthetic control group.

**Main outcomes:** Quarterly counts of vaccinated children following intervention implementation for first doses of the measles, mumps and rubella vaccine (MMR1 at 24 months and at 5 years), second dose of MMR (MMR2 at 5 years), pneumococcal conjugate vaccine (PCV at 24 months), the 6-in-1 vaccine, covering diphtheria, tetanus, pertussis, polio, haemophilus influenzae type b, and hepatitis B (at 12 months), and the rotavirus vaccine (at 12 months).

**Results:** Following HELP, rotavirus vaccine uptake increased by 10.03% (95% CI 0.37% to 24.63%), corresponding to 120 (95% CI 4 to 295) additional infants vaccinated in the intervention group compared to the synthetic control. Similarly, 6-in-1 vaccine uptake rose by 11.56% (95% CI 2.37% to 25.56% ∼143 95% CI 29 to 317 additional children vaccinated. No statistically significant changes were observed for MMR1, MMR2, or PCV. Improvements were short-lived, with uptakes returning to pre-intervention levels after approximately nine months.

**Conclusions:** Community-engagement vaccination interventions may produce a modest short-term improvement in uptake of selected early life vaccines but show limited evidence of benefit for MMR uptake. Our findings suggest that such approaches are unlikely to have a sustained impact without long-term investment, integration into existing immunisation systems and addressing the wider social determinants of health.

**What is already known on this topic?:**

- Childhood vaccination rates in England have declined over the last decade and inequalities in uptake are persistent andwidening.
- Children in socioeconomically deprived areas are less likely to receive routine vaccinations, reflecting both structural barriers and vaccine hesitancy driven by misinformation and lack of trust.
- Innovative community engagement interventions are recommended to address these inequalities, yet evidence of their effectiveness remains limited.

**What this study adds?:**

- Our study shows that hyperlocal community engagement interventions can increase uptake of early-life infant vaccines (rotavirus and 6-in-1) by around 10-12% but provides limited evidence of similar improvements for the MMR vaccine.
- The observed improvements in infant vaccines were transient, returning to baseline levels after approximately nine months, suggesting that one-off initiatives may not produce sustained public health gains without tackling wider social determinants of health.

## Introduction

Childhood vaccinations substantially reduce morbidity and mortality from conditions such as measles and meningitis [1]. To ensure sufficient population-level protection, the World Health Organization (WHO) recommends at least 95% vaccination coverage [2]. However, global vaccination rates for many key vaccines remain below this threshold. In 2024, only 84% of children worldwide received a first dose of measles vaccine by their second birthday, with stagnation and declines for other childhood vaccines continuing since 2019 [2]. Although coverage gaps are more pronounced in low- and middle-income countries, declines have also been observed across 21 of 36 high-income countries [3].

England’s vaccination rates epitomise this global trajectory. Uptake has consistently declined over the past decade and now falls short of the WHO target for every routine childhood vaccination programme [4]. Coverage for the first dose of the measles, mumps, and rubella vaccine (MMR1) reached 88.9% in 2024-2025, the lowest level recorded since 2010-2011 [5]. The public health implications are already surfacing through the resurgence of measles [6–8], with the 2025 Chief Medical Officer (CMO) report signalling an era of frequent outbreaks if coverage is not restored [9]. More importantly, declines are disproportionately affecting children living in poverty and deprived areas. Our previous work shows that the gap in MMR2 uptake at age five exceeded nine percentage points between most and least deprived areas in England in 2019 and continued to widen over time [4], leaving large numbers of the most vulnerable children unprotected. Tackling these widening inequalities has become a global and national priority [10,11].

An important driver of this suboptimal coverage is vaccine hesitancy [12], which the WHO defines as “a delay in acceptance or refusal of vaccines despite availability of vaccination services” [13]. As hesitancy is often driven by misinformation, limited communication, and a lack of trust in health institutions [10], there is a growing need to work with communities with a lower baseline level of confidence to tailor culturally acceptable support [14].

Community-based interventions that are embedded in places where people live, learn, and seek care are recommended and widely implemented to address local concerns, build trust, and improve vaccine confidence [15,16]. In Liverpool (England), the Health Equity Liverpool Project (HELP) serves as an example of a hyperlocal, community engagement intervention implemented to improve MMR uptake among preschool children [17]. Activities were co-developed with communities and local partners and delivered across central and north Liverpool, areas with persistently low uptake. These areas have experienced repeated measles alerts and, more recently, one of the lowest levels of measles protection in England, with only 75.5% of children in Liverpool estimated to be fully protected with two doses of MMR in 2024-2025 [18].

Despite this growing investment in community engagement vaccination programmes, the evidence of their effectiveness remains limited [16]. We used synthetic control methods (SCM) to evaluate the impact of the HELP intervention on uptake of five routine childhood vaccines: MMR1, MMR2, pneumococcal conjugate vaccine booster dose (PCV), the 6-in-1 vaccine (covering diphtheria, tetanus, pertussis, polio, haemophilus influenzae type b, and hepatitis B), and rotavirus vaccine.

## Methods

The study protocol containing details of the public health implications, analytical framework, and potential data sources and indicators has been previously published [19]. We briefly describe the key methodological approach below.

### Intervention

HELP was a community-based initiative implemented across central and north Liverpool between October 2023 and June 2024. The programme aimed to increase childhood vaccination uptake by addressing vaccine hesitancy among parents and carers of children aged 13 months to 5 years and 11 months, with particular emphasis on improving awareness and confidence in the MMR vaccine.

Implementation relied on Community Innovation Teams (CITs), multidisciplinary groups that sit at the interface between communities and formal health services. This included local creatives, community champions, third-sector organisations, and healthcare providers. The CITs followed a three-phase capacity-strengthening model [20]. First, they collected and analysed local insight data on vaccine hesitancy through a survey of 116 parents of unvaccinated children. Second, CIT members received bespoke training aimed at improving confidence and competence in conducting vaccine-related conversations with parents. Third, the teams worked with local creatives to co-design and deliver community engagement activities and communication materials tailored to the barriers identified by the parents themselves.

Intervention activities focused on creating opportunities for parents and carers to engage in conversations about childhood immunisations in supportive environments. Delivery took place through a combination of community outreach events and school-based engagement, where families could discuss vaccination concerns informally with trusted multilingual professionals. Translated information leaflets were developed to support these activities. The flyers used visual illustrations to explain the risks and potential complications associated with measles and were made available in English, Polish, Roma, and Arabic to improve accessibility across diverse communities.

Across the intervention period, the CITs held 22 engagement events across nine delivery sites. In total, 646 conversations about MMR vaccination were recorded with parents during these events. Targeted immunisation reminder text messages were also sent to 1,965 parents of unvaccinated or partially vaccinated children.

### Data

We used general practice (GP) level quarterly vaccine uptake data for all practices in England from the publicly available Cover of Vaccination Evaluated Rapidly (COVER) programme [21]. For each GP, COVER reports the percentage of eligible children who received routine childhood vaccines by certain ages. We extracted 24 consecutive quarters of data from April 2019 to March 2025 (Q1 to Q24). We then derived the count of vaccinated children for each of our outcomes: MMR1 (at 24 months and at 5 years of age), MMR2 (at 5 years), PCV booster dose (at 24 months), 6-in-1 vaccine (at 12 months), and rotavirus vaccines (at 12 months).

We linked COVER data to a set of covariates that reflect the socioeconomic and demographic context and health-seeking behaviour of the GP-registered population, along with structural measures for included practices. These included the total count of children aged 0-4 and the proportion of boys, both derived from the monthly GP registration data [22]. Socioeconomic deprivation was measured using the 2025 English Index of Multiple Deprivation (IMD-2025), a validated composite variable that captures deprivation across seven domains [23]. Ethnicity was derived from the 2021 census and expressed as the proportion of patients identifying as Black (black, black British, black Welsh, Caribbean, or African), Asian (Asian, Asian British, or Asian Welsh), or Mixed (mixed or multiple ethnic groups). We also incorporated measures of practice capacity and quality of care, including the average number of appointments per registered patient and the percentage of the Quality and Outcomes Framework (QOF) achieved. Patient-reported experience measures were obtained from the annual GP Patient Survey [24] and included the proportion of respondents satisfied with their GP services and those reporting caring responsibilities.

The geographic accessibility of practice was proxied using the average straight-line distance between each GP practice and the Lower Super Output Areas (LSOAs) where its registered population lives. As GPs serve patients from multiple LSOAs, Euclidean distances were first calculated between each practice’s physical location and the population-weighted centroid of each LSOA from which the practice had registered patients. These values were then averaged, using LSOA-patient counts as weights, to obtain a single measure for each GP.

### Analysis

We first transformed the quarterly uptake data into a panel format with one observation per GP per quarter. Covariates were transformed to match this quarterly structure as detailed in the study protocol [19]. In brief, monthly variables were averaged across each three-month period, while annually reported covariates were applied uniformly across quarters within the corresponding year. IMD (2025), ethnicity (2021), and the distance measure (calculated at the start of the intervention period) were used as fixed values across all quarters.

Covariates were then linked to the uptake dataset. Variables measured directly at GP level (e.g., child population density and sex distribution, QOF achievement, appointments per patient, patient satisfaction, caring responsibilities, IMD 2025) were linked using the unique GP code. LSOA level variables (ethnicity and patient-weighted average distance to practice) were aggregated to GP level using the GP-LSOA registered population as weights. Detailed data cleaning procedures and a worked example are available in Appendix 1 and on GitHub (https://github.com/Mohammedsherifamin/HELP-evaluation).

GP practices were classified as being in the “intervention group” if the population-weighted centroid of their registered patients fell within a 1km radius of any of the nine HELP delivery sites. This distance corresponds to a 10-to-15-minute walking time for most adults and aligns with the common definition of ‘walkable distance’ [25].

All remaining practices in England formed the donor pool from which the synthetic control group was constructed. Using the Microsynth R package [26], this group was generated as a weighted combination of donor practices that closely match the intervention practices in pre-intervention characteristics and vaccination uptake trends [27]. The resulting weighted group represents the counterfactual; the expected vaccination trajectory in the absence of the intervention.

For each outcome, we estimated the average treatment effect as the post-intervention difference in the number of vaccinated children between the intervention and the synthetic control. The matching model was specified with GP as the unit of analysis and quarter as the time index. Pre-intervention trends in vaccinated counts and their corresponding denominators were included as time-varying matching variables, while remaining covariates were averaged across the pre-intervention period and included as time-invariant variables.

As per the study protocol [19], we first attempted to construct the synthetic control using the full pre-intervention period (Q1-Q18). However, the COVID-19 pandemic substantially disrupted routine vaccination delivery and healthcare utilisation in England [28], leading to instability in pre-intervention trends and poor matching quality between intervention and donor practices. We quantified matching quality using the epsilon statistic, which measures the maximum proportional deviation between treated and synthetic control units after weighting (higher values indicate greater imbalance, with zero indicating an exact match [26,29]). The full period model produced an epsilon of 0.17, indicating that the synthetic control could not be adequately calibrated to match the intervention group across pre-intervention trends and covariates, and was therefore not considered appropriate for inference (Supplementary Figure 1).

Consequently, the primary analysis restricted the pre-intervention period to begin in April 2022 (Q13), the point by which most domestic legal COVID-19 restrictions in England had been lifted and NHS services had returned to normal [30]. It also applied a one-quarter lag ending the pre-period at Q19 (2023 Oct-Dec) to allow for delayed behavioural responses to the intervention (Figure 1). In addition, non-intervention GP practices within Central and North Liverpool primary care networks (PCNs; groups of GP practices that work together) were excluded from the donor pool to minimise potential spillover effects.

**Figure 1.**
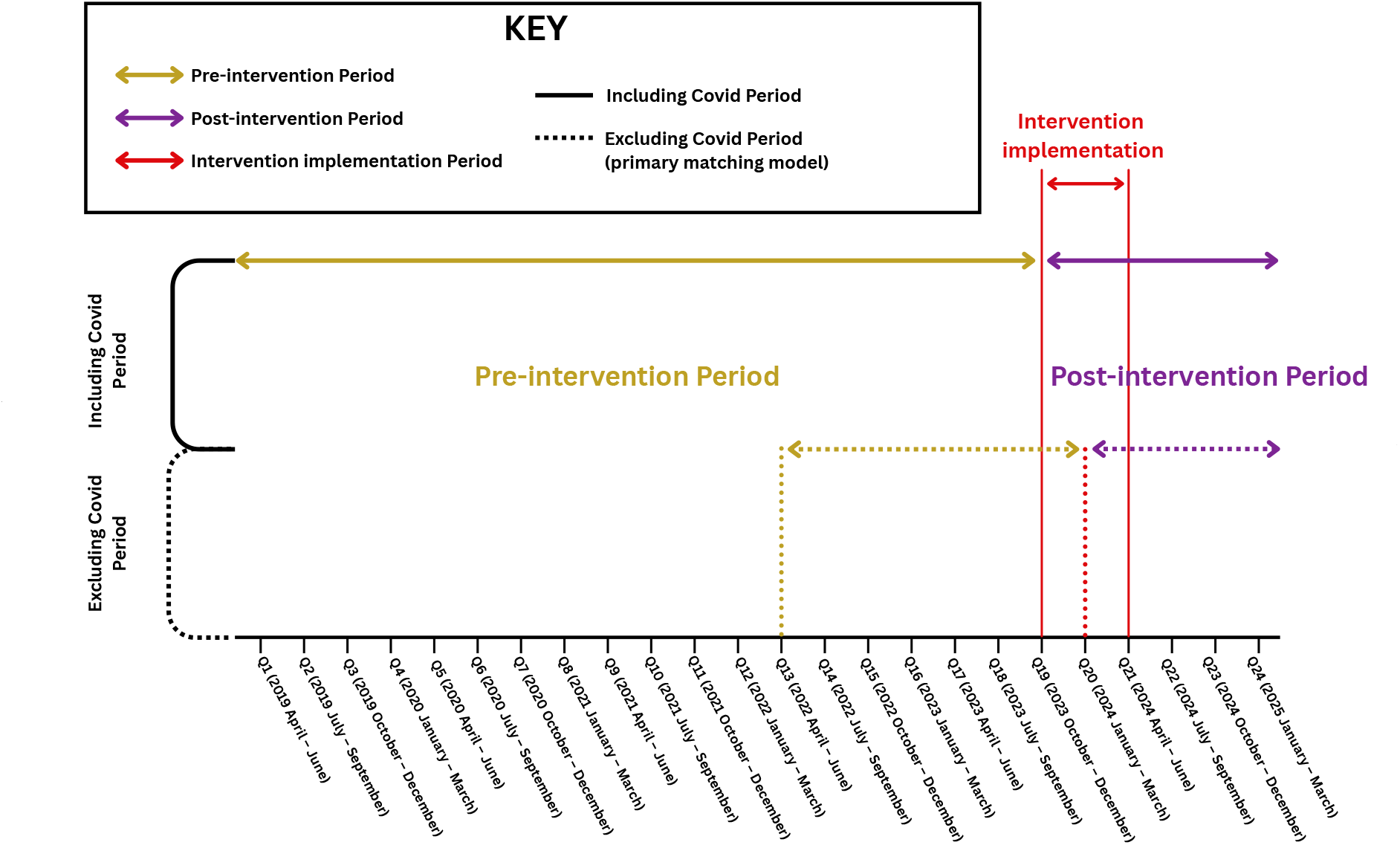
Timeline of the study showing the pre-intervention period, the HELP intervention period, and the post-intervention follow-up.

Statistical significance was assessed using permutation inference with 250 random placebo reassignments of control practices into the intervention practices. Uptake trajectories were displayed with 95% confidence intervals (CI) derived from survey-weighted ratio estimators applied to the microsynth weights.

We also repeated the analysis with different spatial definitions of the intervention group (0.5 km and 1.5 km) to assess model sensitivity and examined matching quality across a wider range of buffer distances (0.5 to 3 km by 0.1 km increments). All analyses were conducted in R (version 4.4.2).

### Patient and Public Involvement

Community members were involved in the development and delivery of the HELP intervention. The project was co-developed with local communities, with activities informed by identified community needs and designed in collaboration with community partners. However, members of the public were not directly involved in the analytical component of this evaluation study. Further details are available in the published HELP report: https://www.lstmed.ac.uk/news-events/blogs/recite-final-help-report. All findings will be shared through local, regional and national events, with public health authorities, GPs, arts organisations, local and national government and communities. Newsletters with lay summaries of the study findings will also be shared publicly.

## Results

### Descriptives

Figure 2 shows the flow diagram of study inclusion. The final analytic sample included 5845 GP practices, contributing a total eligible population of 1,644,027 children at 12 months, 1,688,002 at 24 months, and 1,855,311 at 5 years. Using the 1 km buffer, 19 practices (4,012 children at 12 months, 4,116 at 24 months, and 4,551 at 5 years) defined the intervention group and the remaining 5826 practices contributed to constructing the synthetic control group.

**Figure 2.**
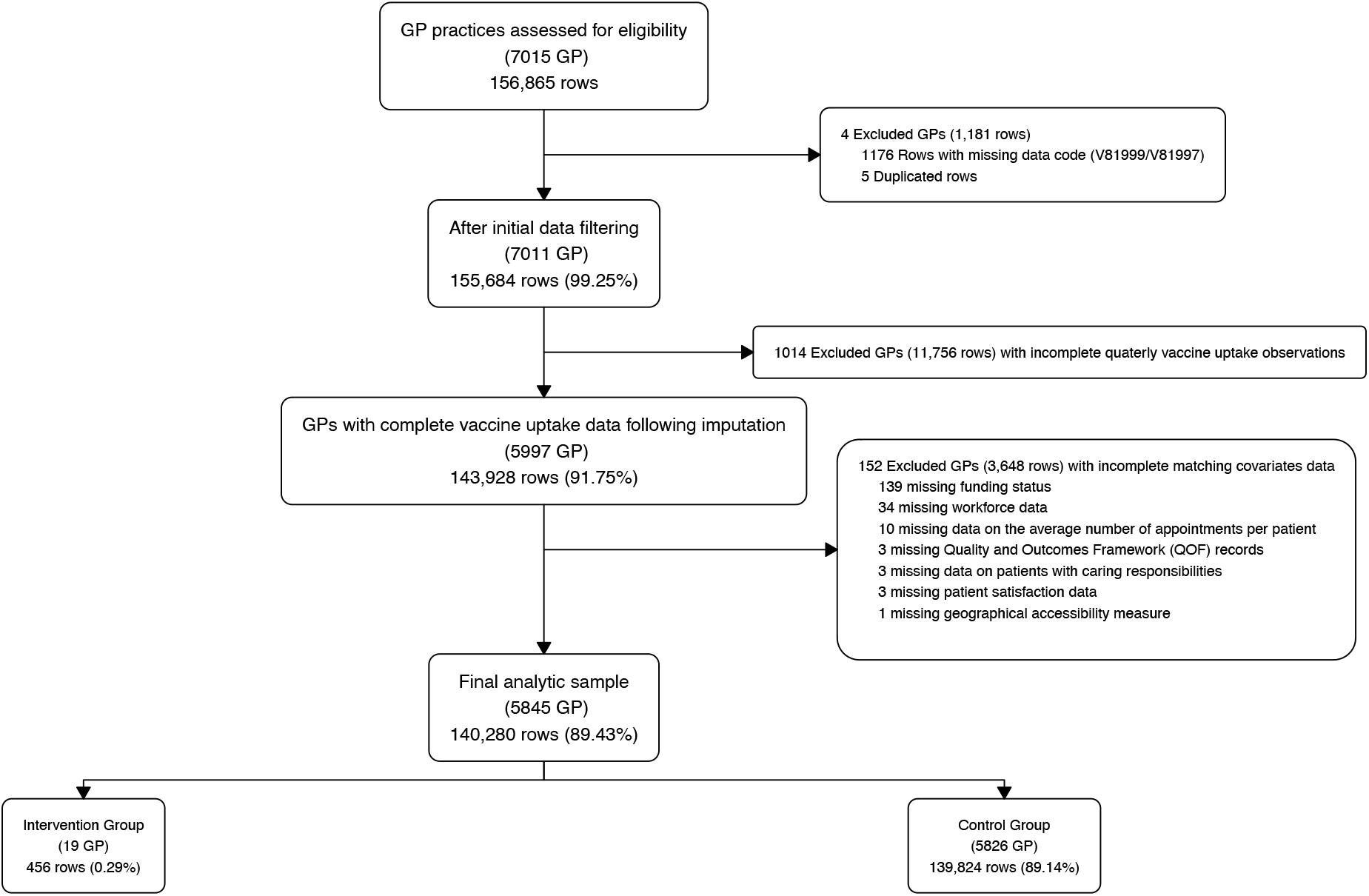
Flow diagram of the study selection showing the final analytic sample and classification into intervention and synthetic control groups based on a 1 km buffer radius around intervention sites.

Table 1 summarises the baseline characteristics during the pre-intervention period (Q13-Q18) prior to the synthetic control construction. The median uptake rates across all vaccines were lower in intervention practices than in non-intervention donor practices. The largest difference was observed for MMR2 at 5 years, with intervention practices averaging 70% compared with 88% in the non-intervention group. A similar pattern was observed for MMR1 at 24 months, rotavirus, and PCV vaccines, where median uptake in intervention practices was lower (78%, 79%, and 78%) compared with non-intervention practices (93%, 92%, and 92%). Smaller differences noted for the remaining vaccines.

**Table 1.** Summary statistics of the pre-intervention period (Q13 - Q18) for the intervention and non-intervention groups before the synthetic control construction.

|  | Intervention GPs<br>(n = 19 practices)<sup>†</sup> | GPs in the rest of England<br>(n = 5826 practices)<sup>‡</sup> |
| --- | --- | --- |
| Eligible population (12 months)* | 1,996 [15 (10, 25)] | 833,732 [20 (12, 30)] |
| Eligible population (24 months)* | 2,058 [16 (11, 25)] | 844,830 [20 (13, 31)] |
| Eligible population (5 years)* | 2,323 [19 (12, 27)] | 934,872 [23 (14, 34)] |
| % MMR1 uptake at 24 months | 78 (67, 87) | 93 (86, 100) |
| % MMR1 uptake at 5 years | 86 (80, 93) | 95 (90, 100) |
| % MMR2 uptake at 5 years | 70 (59, 79) | 88 (80, 95) |
| % 6-in-1 vaccine uptake | 81 (71, 89) | 94 (89, 100) |
| % Rotavirus vaccine uptake | 79 (68, 88) | 92 (85, 97) |
| % PCV booster uptake | 78 (67, 88) | 92 (85, 100) |
| IMD score (2025) | 46 (40, 51) | 21 (14, 31) |
| % Black/Black British | 5.7 (3.0, 8.7) | 1.5 (0.5, 5.9) |
| % Asian/Asian British | 8 (3, 9) | 5 (1, 13) |
| % Mixed ethnicity | 4.27 (2.02, 5.58) | 2.34 (1.38, 3.87) |
| % Other ethnicity | 4.91 (3.28, 7.70) | 1.08 (0.45, 3.10) |
| % Boys aged 0-4 | 51.05 (50.34, 52.01) | 51.26 (50.03, 52.45) |
| Appointments per patient | 5.39 (4.56, 6.60) | 5.35 (4.56, 6.33) |
| QOF score | 89.1 (88.0, 91.0) | 94.3 (91.4, 96.9) |
| % of patients with caring responsibilities | 17.8 (16.1, 20.5) | 18.0 (15.8, 19.9) |
| % Patient satisfaction | 79 (73, 82) | 79 (72, 86) |
| Average distance from GP (km) | 1.23 (1.00, 1.51) | 1.59 (1.21, 2.19) |

*Data is presented as median (25th, 75th percentiles) except for population where the total is presented first, followed by median per practice*
<sup>†</sup>Intervention practices within 1 km of intervention sites
<sup>‡</sup>Non-intervention practices used to generate the synthetic control group
\*Sum across quarters [median (25th, 75th percentiles)]; eligible population counts allow repeat inclusion of the same child across age groups; a child counted at 12 months could be counted again at 24 months and at 5 years.
GP = General Practice; MMR = measles, mumps and rubella vaccine; 6-in-1 = diphtheria, tetanus, pertussis, polio, haemophilus influenzae type b, and hepatitis B; PCV = pneumococcal conjugate vaccine booster dose; IMD = Index of Multiple Deprivation; QOF = Quality and Outcomes Framework.

On average, intervention practices served more socioeconomically disadvantaged populations, reflected by a higher median IMD score (intervention: 46, non-intervention: 21), and had a greater proportion of ethnic minority groups, including Black, Asian, and Mixed ethnicities. The proportions of boys were similar across all groups at 51%. Practice level characteristics and patient-reported measures were also comparable, including the average number of appointments and satisfaction with GP services. However, intervention practices showed slightly lower achievement in the QOF (intervention: 89, non-intervention: 94).

### Statistical analysis

Figure 3 presents the trends in average uptake rates for all outcomes in the intervention and synthetic control groups between April 2022 (Q13) and March 2025 (Q24) under the primary model specification. The model achieved an optimal pre-intervention balance (epsilon = zero), with both groups showing identical uptake trajectories across all vaccines (as well as matched on covariates to have equivalent characterisGcs).

**Figure 3.**
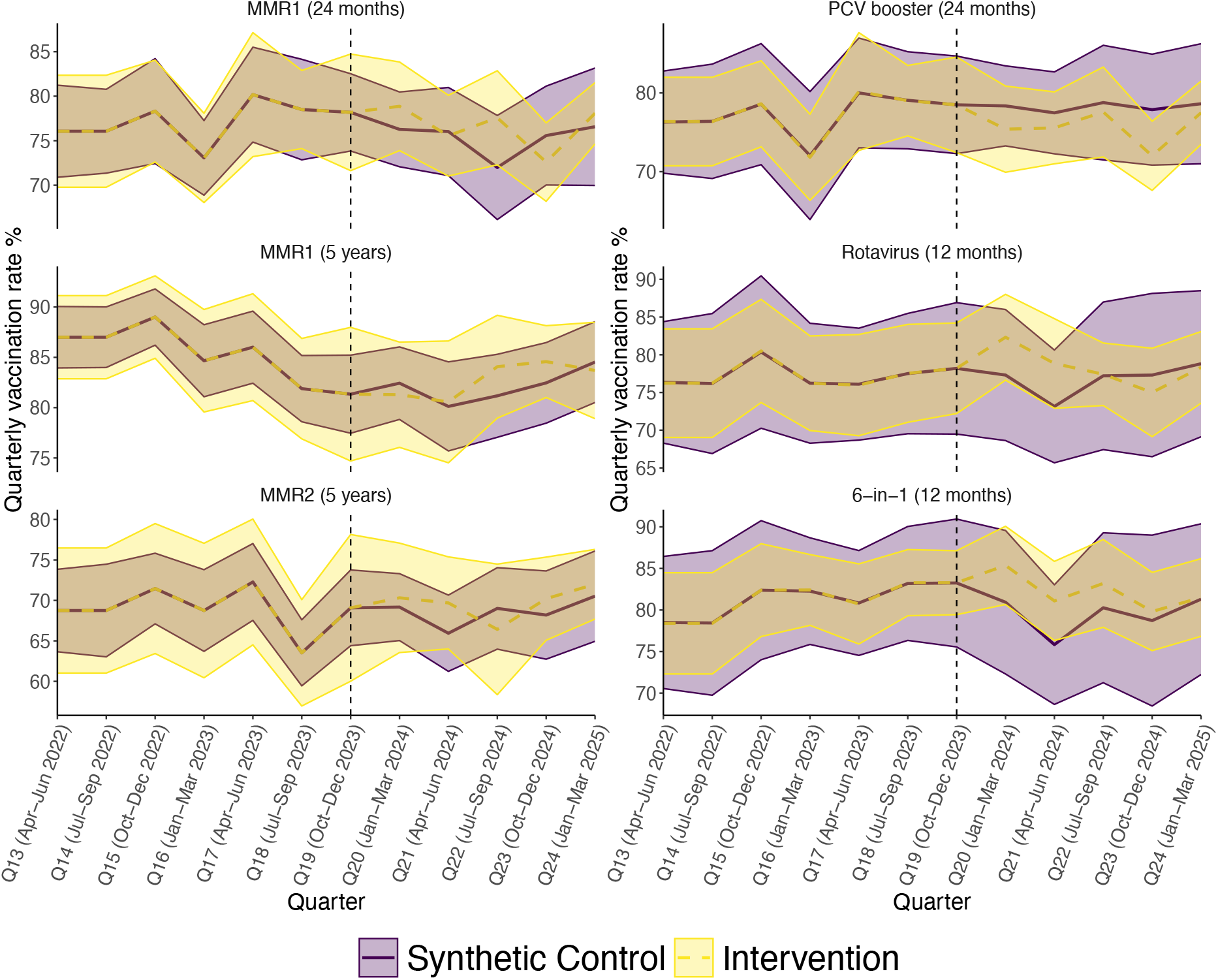
Trend in quarterly uptake rates of MMR1 at 24 months, MMR1 at 5 years, MMR2 at 5 years, rotavirus, 6 in 1, and PCV booster vaccines under the primary matching model. Comparison between the intervention group, defined by a 1km proximity to HELP delivery sites, and the synthetic control group. The black dotted line marks the intervention starting point. Pre-intervention period: Q13 [2022 April-June] to Q19 [2023 October-December]; post-intervention period is from Q20 [2024 January-March] onward; excluding non-intervention practices within Central and North Primary Care Networks.

Following implementation of the HELP intervention, the uptake of the rotavirus vaccine increased by 10.03% (95% CI 0.37% to 24.63%) in the intervention group compared to the synthetic control (Figure 4), corresponding to an estimated 120 additional vaccinated infants (95% CI 4 to 295). This was reflected as an immediate sharp increase in the uptake trajectory, which subsequently moderated over the following nine months (Figure 3). A comparable gain was observed for the 6-in-1 vaccine, where uptake rose by 11.56% (95% CI 2.37% to 25.56%), equivalent to approximately 143 additional vaccinated children (95% CI 29 to 317). In contrast, the intervention did not yield statistically significant changes for MMR1 (at 2 or 5 years), MMR2 (at 5 years), or the PCV booster dose. Figure 5 illustrates the spatial distribution of the non-intervention practices contributing to the synthetic control construction, along with their corresponding matching weights.

**Figure 4.**
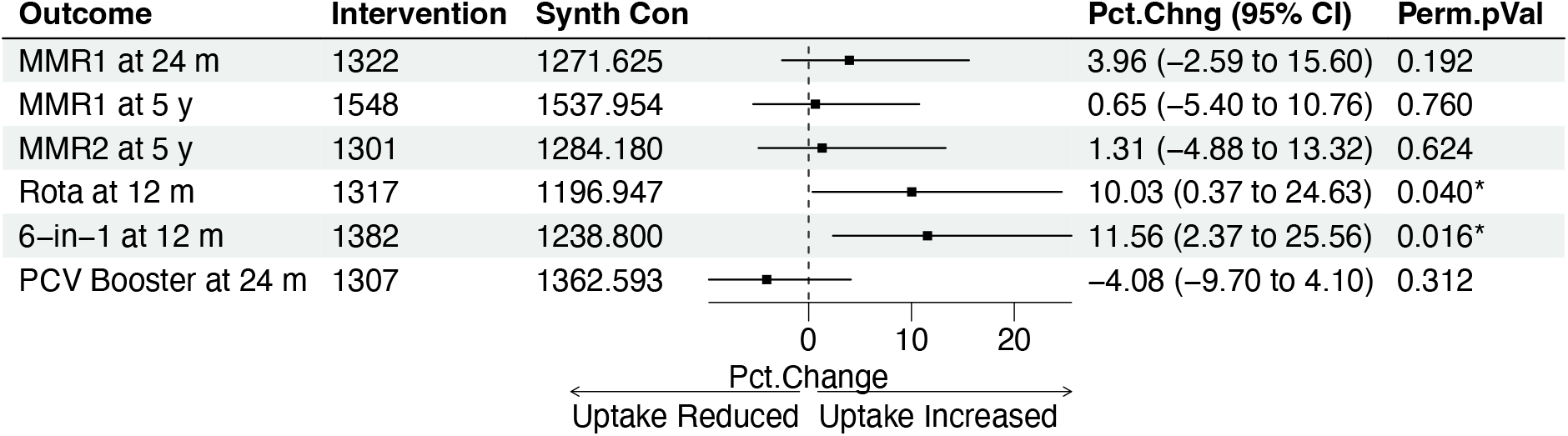
Forest plot of estimated intervention effects in the post-intervention period (Q20 January 2024-Q24 March 2025).

**Figure 5.**
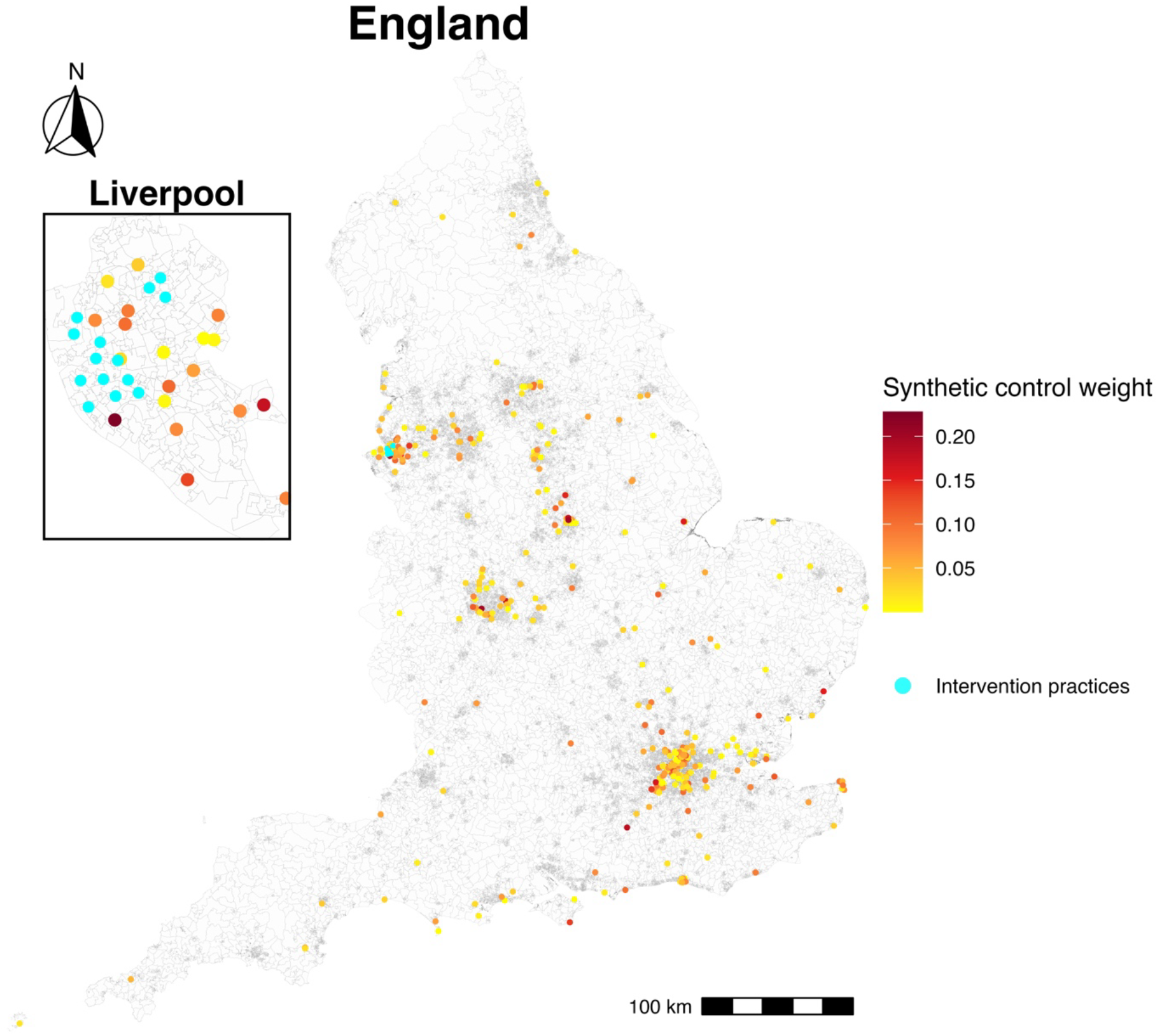
A map of England showing the spatial distribution of non-intervention practices contributing to the construction of a synthetic control group. Non-intervention GPs are shown as coloured dots, with darker shades indicating higher matching weights. Intervention practices are highlighted in cyan.

When we redefined practices’ exposure status using 0.5km and 1.5km proximity from HELP delivery sites, the intervention group included 9 and 34 GPs, respectively (Supplementary Figure 2). The model using the 0.5 km threshold produced poor pre-intervention balance (Supplementary Figure 3) with an epsilon of 0.18 and therefore did not yield reliable estimates. Meanwhile, the 1.5 km specification achieved an exact pre-intervention match (Supplementary Figure 4). Under this specification, significant increases were observed in uptake of both the rotavirus vaccine (8.30%, 95% CI 1.84% to 14.94%) and the 6-in-1 vaccine (7.74%, 95% CI 1.7% to 14.88%) following the intervention implementation (Supplementary Figure 5).

In general, the matching quality across buffer distances showed a sharp decline in imbalance (epsilons) with increasing spatial radius, which subsequently increased the number of intervention units. The 1 km radius was the smallest at which a fully balanced synthetic control could be achieved (Supplementary Table 1, Supplementary Figure 6).

## Discussion

The HELP community-based intervention led to measurable improvements in the uptake of two early childhood vaccines when evaluated using a matched synthetic control constructed from national open-access data. Uptake of the rotavirus vaccine increased by 10.03%, corresponding to approximately 120 additional infants vaccinated, while uptake of the 6-in-1 vaccine increased by 11.56%, equivalent to around 143 additional vaccinated children. Although these absolute numbers are modest, they represent additional protected children in a socioeconomically deprived setting with historically low uptake, where even small gains can be meaningful in reducing susceptibility at population level.

The impact, however, was short-lived, as gains were maintained for approximately nine months before gradually returning to pre-intervention levels, indicating a limited long-term legacy of the programme. This pattern follows previous evidence suggesting that community engagement and behavioural strategies can improve routine vaccination outcomes, but the durability of change requires continued reinforcement rather than one-off implementation [16,31,32]. In the case of HELP, delivery of the intervention was shorter than originally planned. While the programme was initially intended to run from October 2023 to June 2024, most activities were cut short in December 2023 due to funding constraints. This shortened delivery period meant that several planned outputs were not fully realised, which may have limited both the intensity of exposure and the potential durability of the observed effects.

Effect sizes were also attenuated (rotavirus vaccine: 8.30%; 6-in-1 vaccine: 7.74%) when exposure was defined using a wider buffer (1.5 km) around intervention sites, consistent with distance-related decay. As the distance from the intervention sites increases, practices are more likely to be indirectly exposed through shared community networks or diffusion of messaging, which dilutes the observed effect by reducing the differences between intervention and comparison groups [33].

Notably, the intervention primarily targeted MMR uptake, yet we did not observe improvements for MMR outcomes. One plausible explanation is that vaccine behaviours may be easier to influence earlier in infancy, when contact with health services is more frequent and vaccination visits are already scheduled alongside other infant care, creating a built-in reminder system that supports uptake. As children grow older, vaccination becomes more dependent on parental initiation, as is the case with later childhood vaccines such as MMR. This dynamic partially explains the recent changes to the UK childhood immunisation schedule, which moved the timing of the second MMR dose earlier in childhood, from 3 years and 4 months to 18 months [34].

MMR uptake may also be particularly resistant to change because it has long been affected by vaccine safety misinformation, associated with the Andrew Wakefield scandal [35]. Despite extensive evidence from large cohort studies and meta-analyses showing no link between MMR vaccination and autism [36], concerns still shape parental decisions more strongly for MMR than for other routine vaccines [37]. Insights from the parent survey conducted by the CITs during HELP support this interpretation. Parents of unvaccinated children reported concerns about a perceived link between MMR specifically and autism, and some expressed beliefs that the potential risks of vaccination outweighed the risk of measles infection. Concerns were also raised about whether the vaccine had been adequately tested in ethnically diverse populations.

Previous studies evaluating community engagement interventions have generally reported positive effects, with up to a 25% increase in immunisation uptake [38]. Similarly, systematic reviews of behavioural and reminder-based strategies report improvements in uptake, particularly when interventions are endorsed by trusted healthcare providers or community networks [39]. However, much of this evidence originates from low- and middle-income countries [39–41], where baseline vaccination coverage is often lower, and there is a greater margin for improvement.

Evidence from high-income countries remains limited, and much of what exists is derived from randomised control trial settings [32], which may not be directly applicable to population-level community engagement programmes of the kind evaluated in our study. Nonetheless, observational evaluation of large community health worker programmes in the United States has shown approximately a five percentage-point increase in completion of multiple early childhood vaccine series by age five [42], a finding that is less than the magnitude of impact we observed in our study.

### Public health and policy implications

The modest effects observed in our study should be interpreted in the wider context of systemic barriers to vaccine uptake. Structural factors such as poverty [43] and housing [4,44], access [12,45], workforce constraints [46], and long-term underinvestment in preventative services [47] continue to shape vaccination behaviours in deprived communities in ways that local engagement alone cannot overcome. An intervention that reached nearly 2,000 families through reminder messaging and over 600 direct conversations produced modest, time-limited gains in two infant vaccines and no measurable change in MMR uptake, not because community engagement does not work, but because short-term, under-resourced initiatives cannot fully compensate for the effects of deeply rooted structural disadvantages [16]. Our findings are therefore relevant to current policy discussions in England, including the UK parliamentary inquiry into childhood vaccination uptake [48]. We demonstrated the potential and limitations of community engagement approaches when implemented as a one-time, standalone initiative, highlighting the need for parallel investment in addressing the wider social determinants of health and strengthening immunisation systems proportionate to need.

### Strengths and weaknesses

To our knowledge, this is among the first studies in England to apply a synthetic control framework for evaluating a community engagement childhood vaccination intervention, moving beyond uncontrolled before-and-after designs and subjective comparator selection. We considered multiple vaccine outcomes across the childhood vaccination schedule, which allowed us to estimate the differential impact of community-based approaches on early- and later-life vaccination behaviours. We demonstrated the feasibility of using a national open-access dataset, covering all registered practices in England, to conduct a robust quasi-experimental evaluation, without relying on the often access-restricted individual-level data. Importantly, we also addressed major sources of bias that complicate real-world evaluation of vaccination programmes, especially those related to major system shocks, delayed intervention effects, and geographic spillover. Nonetheless, clearer methodological frameworks are needed to guide evaluations of place-based hyperlocal childhood vaccination interventions, particularly under data governance constraints.

Our study still has some limitations. First, the validity of the SCM largely depends on achieving a good pre-intervention match between intervention and comparison units [49]. In our study, matching quality was sensitive by the spatial definition of exposure. The a priori chosen 1 km buffer was the smallest spatial radius at which an exact pre-intervention match was achieved while retaining sufficient contrast to detect intervention effects.

Second, we were unable to examine differential effects of the intervention by child sex due to the aggregated nature of the data, though this is unlikely to affect our findings, as vaccination decisions in early childhood are primarily driven by parental choice rather than child sex. Another limitation was related to the temporal resolution of the data. Outcomes are observed at quarterly intervals, which restricts the precision with which the duration of effects can be estimated. Consequently, the findings should be interpreted as evidence that effects persist for approximately nine months, rather than as a precise estimate of duration.

Third, although SCM strengthens our ability to make robust inferences, the analysis remains observational and cannot rule out residual confounding from unmeasured factors. Local service changes or concurrent initiatives that were not captured may have influenced the uptake independent of the intervention. As outlined in the study protocol [19], we attempted to identify such initiatives through an audit form sent to public health teams. However, the information collected was not sufficiently comprehensive to be incorporated in the analysis and reflects a broader challenge in evaluating hyperlocal public health interventions.

Lastly, our decision to use the matching covariates as averaged values per GP over the study period rather than time-varying measures might have masked any within-practice short term variation. Coefficients of variation indicated minimal temporal variability for most covariates (Supplementary Figure 7). The greater variability observed in the self-reported caring responsibility measure could be due to the known low GP survey response rates [50]. This variable was retained as the best available proxy for parental capacity given data constraints.

## Conclusion

Our study provides empirical evidence on the potential contribution of hyper-local community engagement approaches to improving routine childhood vaccination uptake in settings with persistent coverage gaps. While the HELP programme led to a modest increase in the uptake of some infant vaccines, the absence of measurable change for MMR and the limited duration of the observed improvements highlight the challenges of addressing entrenched barriers to vaccination through short-term interventions alone. These findings suggest that community-based initiatives may play a supportive role in strengthening routine vaccination programmes, particularly where trust, access, or local engagement are barriers. However, they are unlikely to achieve sustained improvements without longer-term implementation, integration with broader vaccination strategies and addressing the wider social determinants of health.

## Statements

### Ethics statement

Ethical approval was not required for this study as it used publicly available aggregate data.

### Contributions

MSA is the lead author and guarantor. Conceptualisation: MAG, NX, DHu, MT; Analytical Methodology: MAG, DHu, NX, MSA; Intervention design and implementation: NE, CT, MT, AI, CH, VD, DHo; Formal analysis: MSA; Visualization: MSA; Data Curation: MSA; Writing – Original Draft: MSA, DHu, MAG, NX; Writing – Review & Editing: all authors; Supervision: DHu; Funding acquisition: MT, DHo, DHu, VD.

## Data availability statement

All data used in the study are openly available online at the Cover of Vaccination Evaluated Rapidly (COVER) programme (https://www.gov.uk/government/collections/vaccine-uptake) and the Department of Health and Social Care Fingertips platform (https://fingertips.phe.org.uk). Further details are available in the study protocol (https://bmjopen.bmj.com/content/16/1/e111500.full).

## Transparency statement

The lead authors (the manuscript’s guarantors) affirm that the manuscript is an honest, accurate, and transparent account of the study being reported; that no important aspects of the study have been omitted; and that any discrepancies from the study as planned (and, if relevant, registered) have been explained.

