## Appendix 1 for "Evaluating the impact of a community-engagement intervention on the uptake of childhood vaccines in England: A synthetic control analysis"

### Appendix 1 Data cleaning

This appendix describes the data cleaning and preparation steps applied to the quarterly GP-level vaccination uptake dataset prior to analysis.

We first aggregated the quarterly uptake data in a panel format with one observation per GP per quarter. Missing uptake or denominator values were imputed as follows: for each GP, missing values at the start (Q1) or end (Q24) were replaced by the nearest observed neighbour (Q2 or Q23, respectively), and internal gaps were filled by carrying forward the previous quarter’s value when both adjacent quarters were observed. Practices with missingness that could not be resolved by this logic (e.g., multiple consecutive missing quarters), and those with all 24 time points missing were then excluded. Non-numeric suppression values in the denominator (e.g., “note 1”, “*”) were replaced with a random count between one and four, reflecting the COVER suppression threshold. When the denominator was zero, the corresponding uptake value was set to 0%.
