## Supplementary Figure 1 for "Evaluating the impact of a community-engagement intervention on the uptake of childhood vaccines in England: A synthetic control analysis"

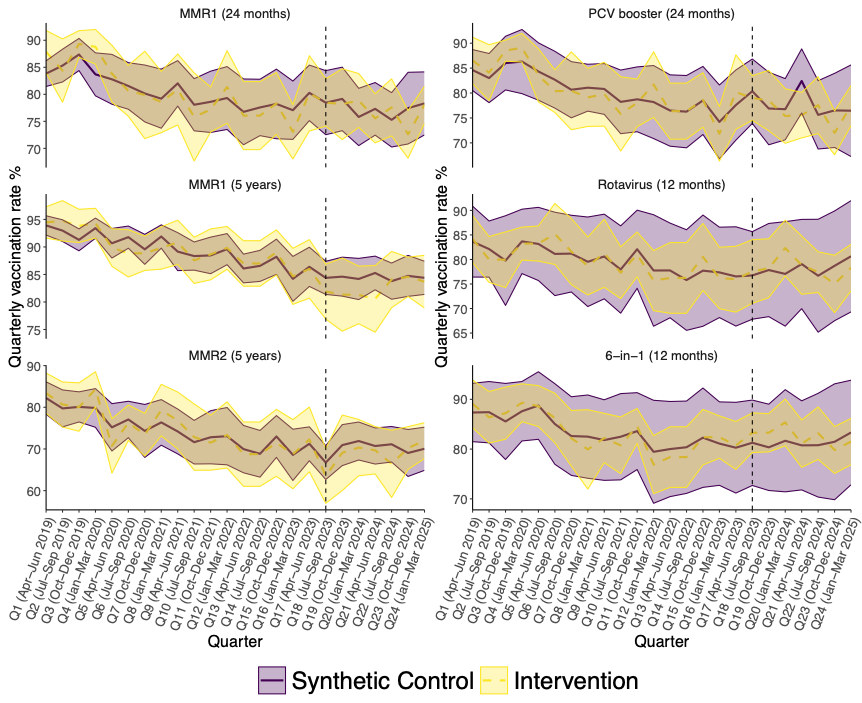

Supplementary Figure 1 Trend in quarterly uptake rates of MMR1 at 24 months, MMR1 at 5 years, MMR2 at 5 years, rotavirus, 6 in 1, and PCV booster vaccines using 1 km buffer distance comparing the intervention and synthetic control groups. The black dotted line represents the start of the intervention. Pre-intervention period: Q1 [2019 April-June] to Q18 [2023 July-September]; post-intervention period is from Q19 [2023 October-December] onward.
