## Supplementary Figure 2 for "Evaluating the impact of a community-engagement intervention on the uptake of childhood vaccines in England: A synthetic control analysis"

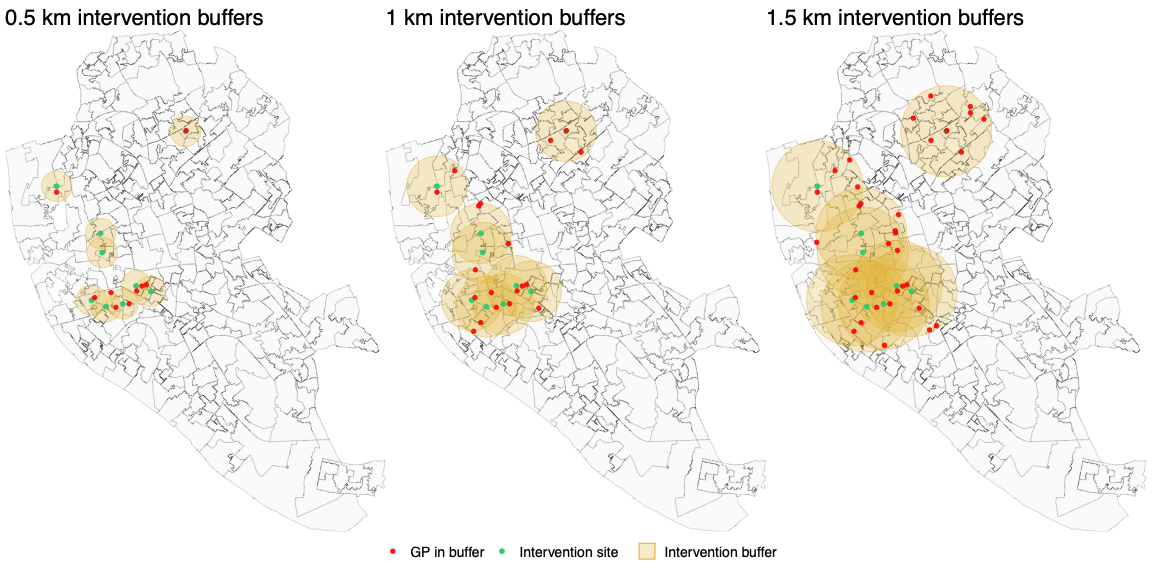

Supplementary Figure 2 Map of Liverpool showing buffer distances around intervention sites. Green dots with yellow rings indicate the locations of intervention sites and their surrounding buffer zones. Red dots represent the population-weighted centroids of GP practices that fall within each buffer distance.
