## Supplementary Figure 3 for "Evaluating the impact of a community-engagement intervention on the uptake of childhood vaccines in England: A synthetic control analysis"

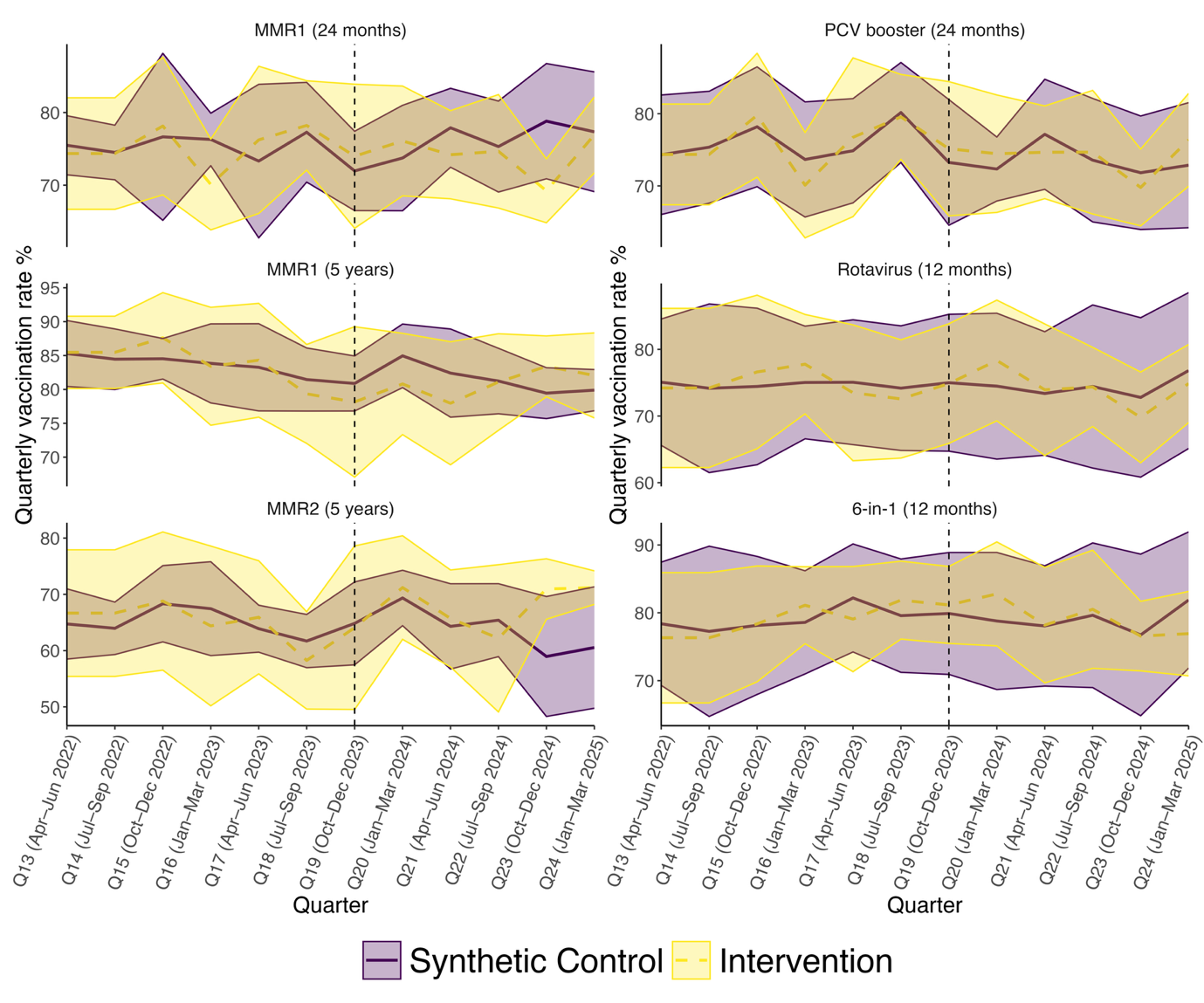

Supplementary Figure 3 Trend in quarterly uptake rates of MMR1 at 24 months, MMR1 at 5 years, MMR2 at 5 years, rotavirus, 6 in 1, and PCV booster vaccines under the primary matching model. Comparison between the intervention group, defined by a 0.5km proximity to HELP delivery sites, and the synthetic control group. The black dotted line marks the intervention starting point. Pre-intervention period: Q13 [2022 April-June] to Q19 [2023 October-December]; post-intervention period is from Q20 [2024 January-March] onward; excluded non-intervention practices within Central and North Primary Care Networks.
