## Supplementary Figure 5 for "Evaluating the impact of a community-engagement intervention on the uptake of childhood vaccines in England: A synthetic control analysis"

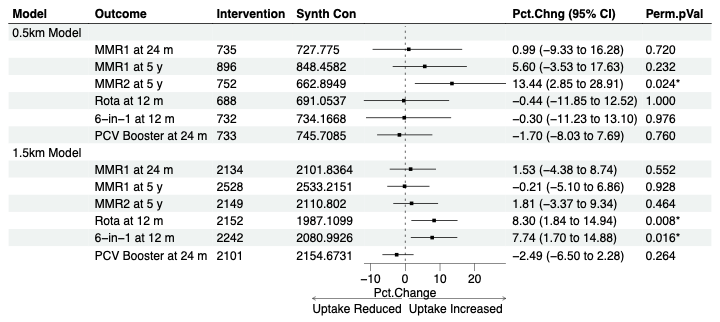

Supplementary Figure 5 Forest plot of estimated intervention effects in the post-intervention period (Q20 January 2024-Q24 March 2025) using a 0.5 and 1.5 km buffer distance.
