## Supplementary Figure 6 for "Evaluating the impact of a community-engagement intervention on the uptake of childhood vaccines in England: A synthetic control analysis"

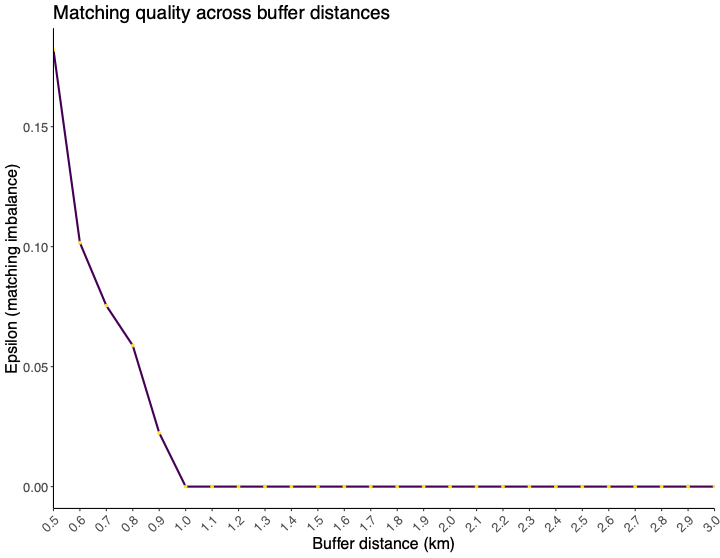

Supplementary Figure 6 Matching quality (epsilon) across buffer distances. Epsilon values are shown for radii from 0.5 km to 3 km, with lower values indicating improved pre-intervention balance between intervention and synthetic control groups
