## Supplementary Figure 7 for "Evaluating the impact of a community-engagement intervention on the uptake of childhood vaccines in England: A synthetic control analysis"

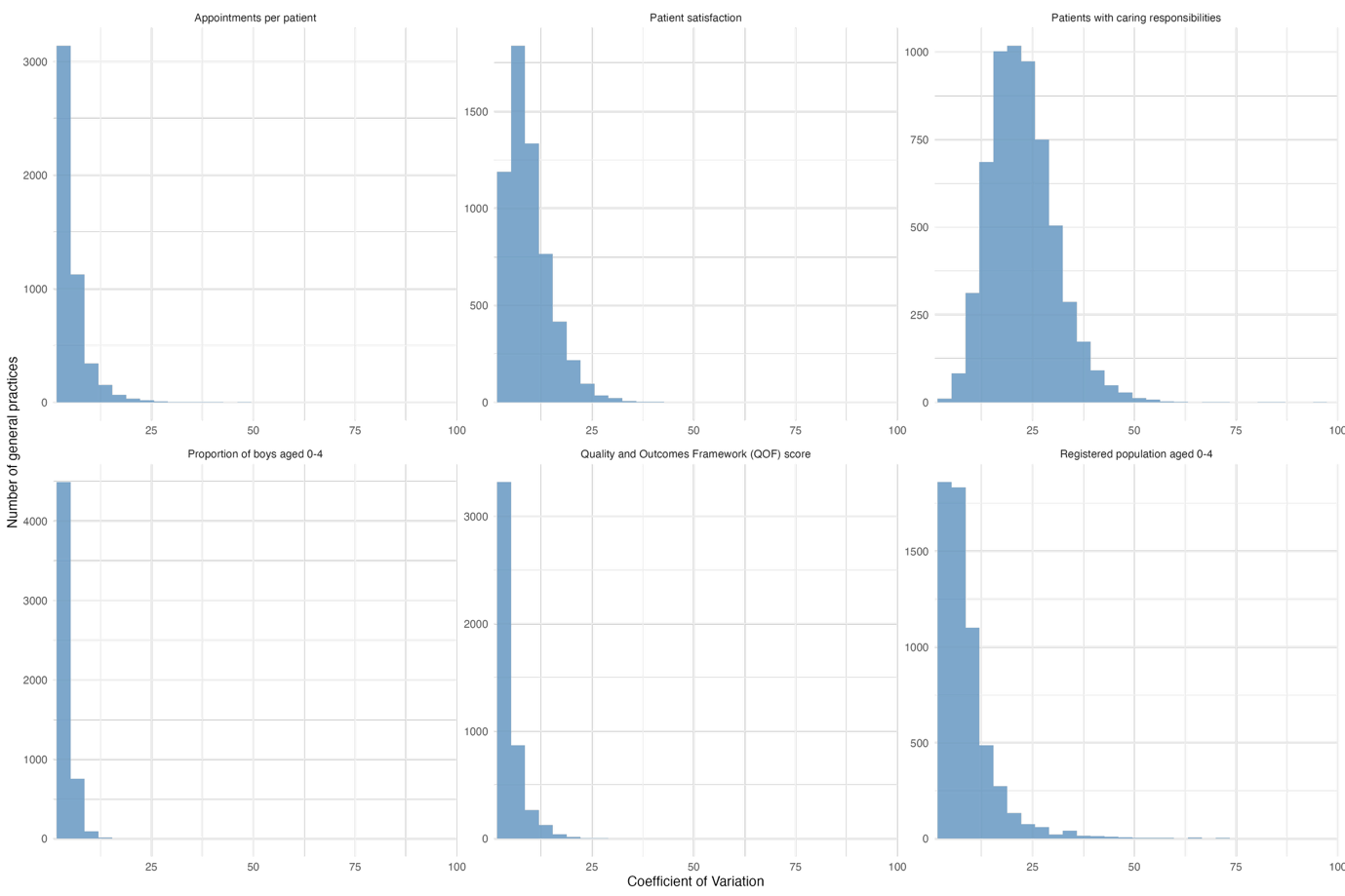

Supplementary Figure 7 Distribution of coefficients of variation for matching covariates across GP practices. Coefficients of variation were calculated at the GP-practice level for each matching covariate as the standard deviation divided by the mean value across the study period and expressed as a percentage
