## Supplementary Table 1 for "Evaluating the impact of a community-engagement intervention on the uptake of childhood vaccines in England: A synthetic control analysis"

Supplementary Table 1 Number of GP practices classified as intervention units and corresponding epsilon values across alternative buffer radii.

| Buffer Radius | Number of Intervention GPs | epsilon |
| --- | --- | --- |
| 0.5 | 9 | 0.18195452 |
| 0.6 | 11 | 0.10163064 |
| 0.7 | 13 | 0.07540313 |
| 0.8 | 14 | 0.05878262 |
| 0.9 | 16 | 0.02237935 |
| 1.0 | 19 | 0.00000000 |
| 1.1 | 20 | 0.00000000 |
| 1.2 | 25 | 0.00000000 |
| 1.3 | 26 | 0.00000000 |
| 1.4 | 31 | 0.00000000 |
| 1.5 | 34 | 0.00000000 |
| 1.6 | 39 | 0.00000000 |
| 1.7 | 40 | 0.00000000 |
| 1.8 | 41 | 0.00000000 |
| 1.9 | 41 | 0.00000000 |
| 2.0 | 43 | 0.00000000 |
| 2.1 | 45 | 0.00000000 |
| 2.2 | 48 | 0.00000000 |
| 2.3 | 50 | 0.00000000 |
| 2.4 | 53 | 0.00000000 |
| 2.5 | 55 | 0.00000000 |
| 2.6 | 56 | 0.00000000 |
| 2.7 | 58 | 0.00000000 |
| 2.8 | 61 | 0.00000000 |
| 2.9 | 64 | 0.00000000 |
| 3.0 | 64 | 0.00000000 |
